# Nearly half of adults with symptoms of sexually transmitted infections (STIs) did not seek clinical care: A population-based study of treatment-seeking behavior among adults in Rakai, Uganda

**DOI:** 10.1101/2023.02.01.23285331

**Authors:** Yasmin P. Ogale, Caitlin E. Kennedy, Fred Nalugoda, Josephine Mpagazi, Jade C. Jackson, Ronald Galiwango, Robert Ssekubugu, Godfrey Kigozi, Julie A. Denison, Charlotte A. Gaydos, Joseph Kagaayi, M. Kathryn Grabowski

## Abstract

Understanding treatment-seeking behavior is critical to the treatment and control of sexually transmitted infections (STIs), yet current data on STI treatment-seeking in low-resource settings is scarce. This study aims to describe STI treatment-seeking behavior and identify factors associated with seeking treatment at a clinic among adults with STI-related symptoms in rural Uganda. The STI prevalence study (STIPS) conducted a population-based survey and STI testing among all consenting adults aged 18–49 in two communities in rural south-central Uganda. Of 1,825 participants, 962 individuals self-reported STI symptoms in the past six months; we present descriptive data on treatment-seeking and STI prevalence among these individuals. We used multivariable Poisson regressions with robust variance to determine the sociodemographic and symptom-related factors independently associated with seeking STI treatment at a clinic and assessed the association with previous clinic treatment-seeking and current STI diagnosis. Forty-three percent of adults who reported STI-related symptoms in the past six months said they did not seek any treatment. Among those who did, 58% sought treatment at a private clinic, 28% at a government clinic, 9% at a pharmacy/drug store, 3% at a traditional healer, 2% at a market/shop, and 5% at another location. Among both men and women, having multiple STI related symptoms was positively associated with clinic treatment seeking (men=PRR: 1.73, 95%CI: 1.36-2.21; women=PR: 1.41, 95%CI: 1.12-1.78). Approximately one-third of men and women who reported previously seeking clinic treatment for their symptoms were diagnosed with a curable STI at the time of the survey. In this setting, nearly half of adults with STI-related symptoms are not seeking clinical care and many who report having sought treatment recent STI symptoms have curable STIs. Future studies should explore barriers to care-seeking and strategies to improve STI services.

## Introduction

Understanding STI treatment-seeking behavior is critical to STI control; timely and appropriate STI treatment can reduce the duration of infectiousness and limit the spread of infection [1, 2]. Treatment-seeking behavior can be defined as any activity undertaken by an individual who perceives themselves to be ill or have a health problem, in order to find a remedy. Various factors can influence the decision to seek treatment, including individual-level factors such as symptom recognition, preexisting beliefs and meanings of symptoms, and perceived efficacy of different treatment methods [1, 2]. Contextual factors such as the availability, quality and accessibility of treatment, as well as social stigma, also play a role [1, 2]. These factors influence not only the timing and location of treatment-seeking, but also whether an individual seeks treatment at all [1, 2].

Despite its importance, population-based data on treatment-seeking in low-resource settings, such as Uganda, is scarce. In such settings, syndromic management by a healthcare worker is used to diagnose presumptive infection [3, 4], and so seeking clinical care is the best way to receive effective treatment. Data from the 2016 Uganda DHS show that, of those who reported having an STI or STI symptoms, 71% of women and 64% of men sought advice or treatment from a clinic, hospital, private doctor or other health professional, while 26% of women and 34% of men did not seek any advice or treatment [5]. Previous studies on STI treatment-seeking behavior in Uganda report a variety of treatment approaches, including public health facilities, private practitioners, traditional healers and self-treatment [6, 7]. A qualitative study from 1999 on STI treatment-seeking behaviors among Ugandan adults found that treatment in the informal sector, including self-treatment and traditional healers, was especially common (over 60% of participants). The study also found that for participants with STI-related symptoms, deterrents to seeking care at public health facilities included long waiting times, lack of drugs, user fees, corruption and bribes by health workers, health workers abusing STI patients, lack of privacy, long distances, fear of being tested for AIDS, specimens not being examined in the laboratory and being given tablets instead of injections (which were preferred) [6]. Previous studies in the region have also found gender differences in treatment-seeking, with barriers to care including lack of access to facilities, lack of time and money, and dependence on men for permission to leave the home, resulting in women either ignoring their problem, using self-care or self-medication, using herbal or traditional medicine, or using services in secret [8–13]. Consultations with husbands, relatives and friends would also delay treatment-seeking for women, with prompt care triggered by symptoms that were perceived to be more severe or life threatening [14].

While useful in providing context, previous studies from low-resource settings, including those mentioned above, are limited in that many sampled participants at treatment facilities only after they presented for care; most studies do not capture individuals who delay seeking treatment or who do not seek treatment at all. This study uses a population-based sample to describe treatment-seeking behavior among adults with

STI-related symptoms in rural Uganda and identify factors associated with seeking treatment at a clinic.

## Methods

### Study Setting

This study was conducted as part of the STI prevalence study (STIPS), a population-based survey conducted in the Rakai region of South-central Uganda that aimed to estimate the population STI burden [15]. STIPS was conducted as a part of the Rakai Community Cohort Study (RCCS), one of the oldest population-based studies of HIV in Africa. Conducted by the Rakai Health Sciences Program (RHSP), the RCCS is an ongoing, open community-based cohort of residents aged 15-49 years in agrarian communities, semi-urban trading centers and Lake Victoria fishing communities in the Rakai region. The RCCS includes the administration of a demographic and health questionnaire, as well as HIV testing for all consenting participants. Details of the RCCS study design can be found elsewhere [16].

STIPS was approved by the Uganda Virus Research Institute Research Ethics Committee (GC/127/19/03/709) and the Johns Hopkins School of Medicine Institutional Review Board (IRB00204691). The study was also registered with the Ugandan National Council for Science and Technology (HS 364 ES).

### Data Collection

STIPS was a population-based survey that recruited all individuals aged 18-49 years from two RCCS communities – one inland and one fishing – from May to October 2019 [15]. Three adjacent villages consisting of semi-urban and rural agrarian populations comprised the inland community and one fish landing site along Lake Victoria comprised the fishing community. In addition to the standard RCCS questionnaire, participants were administered an STI module that assessed their symptom status and treatment-seeking behavior. To ascertain symptom status, each participant was prompted on a list of symptoms and asked to identify each symptom that they had experienced in the past six months (previous symptoms) and also in the past 7 days (recent symptoms). Symptoms included: genital ulcer, genital discharge, frequent urination, painful urination, pain during intercourse, bleeding during intercourse, lower abdominal pain and genital warts, as well as thick and/or colored vaginal discharge, itching of the vagina and unpleasant vaginal odor for women. Treatment-seeking behavior was defined as self-reported treatment-seeking from any location for STI-related symptoms in the past six months, and was assessed among participants who reported experiencing previous or recent symptoms: participants were asked if they did anything to help cure their symptoms or to prevent passing on infection to their spouse or partner(s), and if so, what action(s) did they take: used condoms, abstinence, sought treatment for self, sought treatment for partner or some other action. Those who reported seeking treatment for themselves were also asked to specify where they went for treatment: pharmacy/drug store, market/shop, Rakai Program Clinic (RHSP-run clinic providing HIV care and related services), government doctor/nurse/clinic, private doctor/nurse/clinic, traditional healer or other. Up to three locations were recorded per participant.

In addition to the routine HIV screening conducted in the RCCS, STIPS participants were also evaluated for Chlamydia trachomatis (CT), Neisseria gonorrhoeae (NG), Trichomonas vaginalis (TV) and syphilis. All consenting participants provided genital swabs at the time of interview for testing (clinician-collected penile urethral meatus swabs for men and self-administered vaginal swabs for women). CT/NG testing was performed using the Abbott RealTime CT/NG assay using the Abbott m2000 RealTime System for PCR testing at the RHSP central laboratory. TV testing was performed using the OSOM Trichomonas Rapid Test (Sekisui) at the time of the survey. Syphilis screening was performed using the SD Bioline 3.0, a solid phase immunochromatographic assay for the qualitative detection of antibodies of all isotypes (IgG, IgM, IgA) against T. pallidum. Syphilis screening was performed with HIV testing at time of survey; the rapid plasma reagin test (RPR) was then performed within 24 hours at the RHSP central laboratory for all participants with positive screening results to determine syphilis titers. All assays were conducted according to the manufacturers’ protocol. All individuals who tested positive for any STI were provided treatment by RHSP per the Ugandan National Clinical Treatment Guidelines for STIs.

### Data Analysis

We first measured the overall prevalence of STI-related symptoms among all STIPS participants. For the remaining analyses, we restricted our sample to only those who reported any STI-related symptoms in the past six months (n=962). First, we conducted a descriptive analysis of the data, including assessing the prevalence of previous STI symptoms in the past six months. Second, we measured the prevalence of treatment-seeking overall and at each treatment location (e.g. government clinic, private clinic, pharmacy/drug store) and compared the prevalence of private versus government clinic treatment-seeking. Third, we compared sociodemographic and symptom-related factors among participants who sought treatment at a government or private clinic (clinic) compared to those who sought treatment at a non-clinic location (e.g. pharmacy/drug store, market/shop, traditional healer, etc. or who sought no treatment at all. Finally, we compared the prevalence of any curable STI (NG, CT, TV or active syphilis) between participants who did and did not previously seek clinic treatment, and assessed the univariable association between previous clinic treatment-seeking and current prevalence of curable STIs. All prevalence risk ratios were estimated with modified Poisson regression models with robust variance [17]. Given the different social and economic contexts of men and women, we conducted analyses for the sample as a whole, as well as for each gender separately. We also stratified data for each gender by community type.

Complete treatment-seeking information (i.e. any treatment-seeking [yes or no] and specific treatment location) was collected for 99.4% of the sample, with only 6 participants dropped from the analysis because of missing treatment data. Age was analyzed in five-year age groupings. We also calculated the number of symptoms in the past week, and in the past six months, as a sum of a participant’s self-reported symptoms in the respective time period. We assessed positive STI diagnoses with the STIPS test result, with syphilis RPR titers ≥8 considered indicative of high titer syphilis infection [18].

We used prior information from the literature to critically evaluate and select variables for inclusion in the final multivariable models. Based on a conceptual framework of treatment-seeking behavior [1], we included in our model the number of STI-related symptoms experienced in the past six months, as well as select sociodemographic characteristics (age, community type, marital status, HIV status) that could theoretically affect treatment-seeking behavior. All data analysis was carried out in STATA version 15 [19].

## Results

### Sociodemographic characteristics, sexual behavior and STI symptomatology

Fifty-three percent (962/1,825) of adults reported any STI symptoms in the six months prior to the STIPS interview date (34% [290/860] of males; 70% [672/964] of females). Sixty-three percent (605/962) of those with symptoms in the past six months reported at least one symptom in the past seven days (51% [149/290] of males; 68% [456/672] of females). Table 1 summarizes the demographic characteristics of the study group. Most men were aged 30-39 years, in a monogamous marriage, from the fishing community, Christian, educated at some level and working in the fishing industry. Most women were aged 20-29 years, in a monogamous marriage, from the fishing community, Christian, educated at some level and working in agricultural or housework. Approximately one-third of the study sample were people living with HIV (PLHIV). With respect to sexual behaviors, approximately half of the men in the sample reported 2-4 sexual partners in the past year (148 [51%]) and the majority reported 5-10 sexual partners in their lifetime (205 [71%]). Over three-quarters of women in the sample reported having one sexual partner in the past year (529 [79%]) and just over half of women reported 2-4 lifetime sexual partners (408 [61%]). For unmarried men, the majority reported inconsistent condom use in the past year (48/87 [55%]) while the majority of unmarried women reported never using a condom (90/158 [57%]).

**Table 1.** Sociodemographic characteristics and symptomatology of STIPS participants who reported STI symptoms in the past 6 months (N=962), by gender. Data are presented as n (%).

|  | Total<br>N=962 | Male<br>N=290 | Female<br>N=672 |
| --- | --- | --- | --- |
| Age |  |  |  |
| 15-19 years | 69 (7%) | 14 (5%) | 55 (8%) |
| 20-29 years | 371 (39%) | 98 (34%) | 273 (41%) |
| 30-39 years | 356 (37%) | 115 (40%) | 241 (36%) |
| 40-49 years | 166 (17%) | 63 (22%) | 103 (15%) |
| Marital status |  |  |  |
| Never Married | 75 (8%) | 33 (11%) | 42 (6%) |
| Married, Monogamous | 560 (58%) | 174 (60%) | 386 (57%) |
| Married, Polygamous | 128 (13%) | 23 (8%) | 105 (16%) |
| Previously Married | 199 (21%) | 60 (21%) | 139 (21%) |
| Community type |  |  |  |
| Inland | 420 (44%) | 96 (33%) | 324 (48%) |
| Fishing | 542 (56%) | 194 (67%) | 348 (52%) |
| Religion (N=935) |  |  |  |
| Christian | 806 (86%) | 244 (84%) | 562 (87%) |
| Muslim | 123 (13%) | 45 (16%) | 78 (12%) |
| Other/none | 6 (1%) | 1 (0%) | 5 (1%) |
| Education |  |  |  |
| No | 62 (6%) | 21 (7%) | 41 (6%) |
| Yes | 900 (94%) | 269 (93%) | 631 (94%) |
| Occupation |  |  |  |
| Agricultural or housework | 342 (36%) | 50 (17%) | 292 (43%) |
| Bar or restaurant work | 71 (7%) | 1 (0%) | 70 (10%) |
| Boda boda driving or trucking | 20 (2%) | 20 (7%) | 0 (0%) |
| Fishing | 128 (13%) | 128 (44%) | 0 (0%) |
| Student | 12 (1%) | 4 (1%) | 8 (1%) |
| Trader or shopkeeper | 220 (23%) | 36 (12%) | 184 (27%) |
| Other | 169 (18%) | 51 (18%) | 118 (18%) |
| HIV status (N=960) |  |  |  |
| Negative | 649 (68%) | 200 (69%) | 449 (67%) |
| Positive | 311 (32%) | 90 (31%) | 221 (33%) |
| Sex in the past year |  |  |  |
| No | 55 (6%) | 9 (3%) | 46 (7%) |
| Yes | 907 (94%) | 281 (97%) | 626 (93%) |
| Sexual partners in the past year |  |  |  |
| None | 55 (6%) | 9 (3%) | 46 (7%) |
| 1 | 631 (66%) | 102 (35%) | 529 (79%) |
| 2-4 | 243 (25%) | 148 (51%) | 95 (14%) |
| 5-10 | 25 (3%) | 24 (8%) | 1 (0%) |
| >10 | 8 (1%) | 7 (2%) | 1 (0%) |
| Sex with partner from outside the community |  |  |  |
| No | 711 (74%) | 162 (56%) | 549 (82%) |
| Yes | 251 (26%) | 128 (44%) | 123 (18%) |
| Lifetime sexual partners |  |  |  |
| None | 10 (1%) | 2 (1%) | 8 (1%) |
| 1 | 77 (8%) | 1 (0%) | 76 (11%) |
| 2-4 | 460 (48%) | 52 (18%) | 408 (61%) |
| 5-10 | 376 (39%) | 205 (71%) | 171 (25%) |
| >10 | 39 (4%) | 30 (10%) | 9 (1%) |
| Condom use in past 12 months by marital status |  |  |  |
| Married | 688 (72%) | 197 (68%) | 491 (73%) |
| Not married, never used condoms | 120 (12%) | 30 (10%) | 90 (13%) |
| Not married, sometimes/inconsistent use | 103 (11%) | 48 (17%) | 55 (8%) |
| Not married, always used condoms | 22 (2%) | 9 (3%) | 13 (2%) |
| NA, no sex | 29 (3%) | 6 (2%) | 23 (3%) |
| Symptoms in the past 7 days |  |  |  |
| No | 357 (37%) | 141 (49%) | 216 (32%) |
| Yes | 605 (63%) | 149 (51%) | 456 (68%) |
| Number of STI symptoms in past 6 months |  |  |  |
| 1 | 313 (33%) | 161 (56%) | 152 (23%) |
| 2-4 | 462 (48%) | 120 (41%) | 342 (51%) |
| >=5 | 187 (19%) | 9 (3%) | 178 (26%) |
| Number of STI symptoms in past 7 days |  |  |  |
| 0 | 357 (37%) | 141 (49%) | 216 (32%) |
| 1 | 261 (27%) | 101 (35%) | 160 (24%) |
| 2-4 | 250 (26%) | 42 (14%) | 208 (31%) |
| >=5 | 94 (10%) | 6 (2%) | 88 (13%) |

In terms of symptoms, the majority of men reported only one symptom (161 [56%]) in the past six months, with painful urination being the most common (42%). The majority of women reported 2-4 symptoms (342 [51%]) in the past six months, with vaginal itching being the most common (62%) (Figure 1). Half (149 [51%]) of men reported experiencing symptoms in the seven days before the interview as compared to 68% (456) of women. Of those who reported symptoms in the past seven days, most men reported one symptom in the past week (101 of 149 [68%]) and most women reported 2-4 symptoms in the past week (208/456 [46%]).

**Fig 1.**
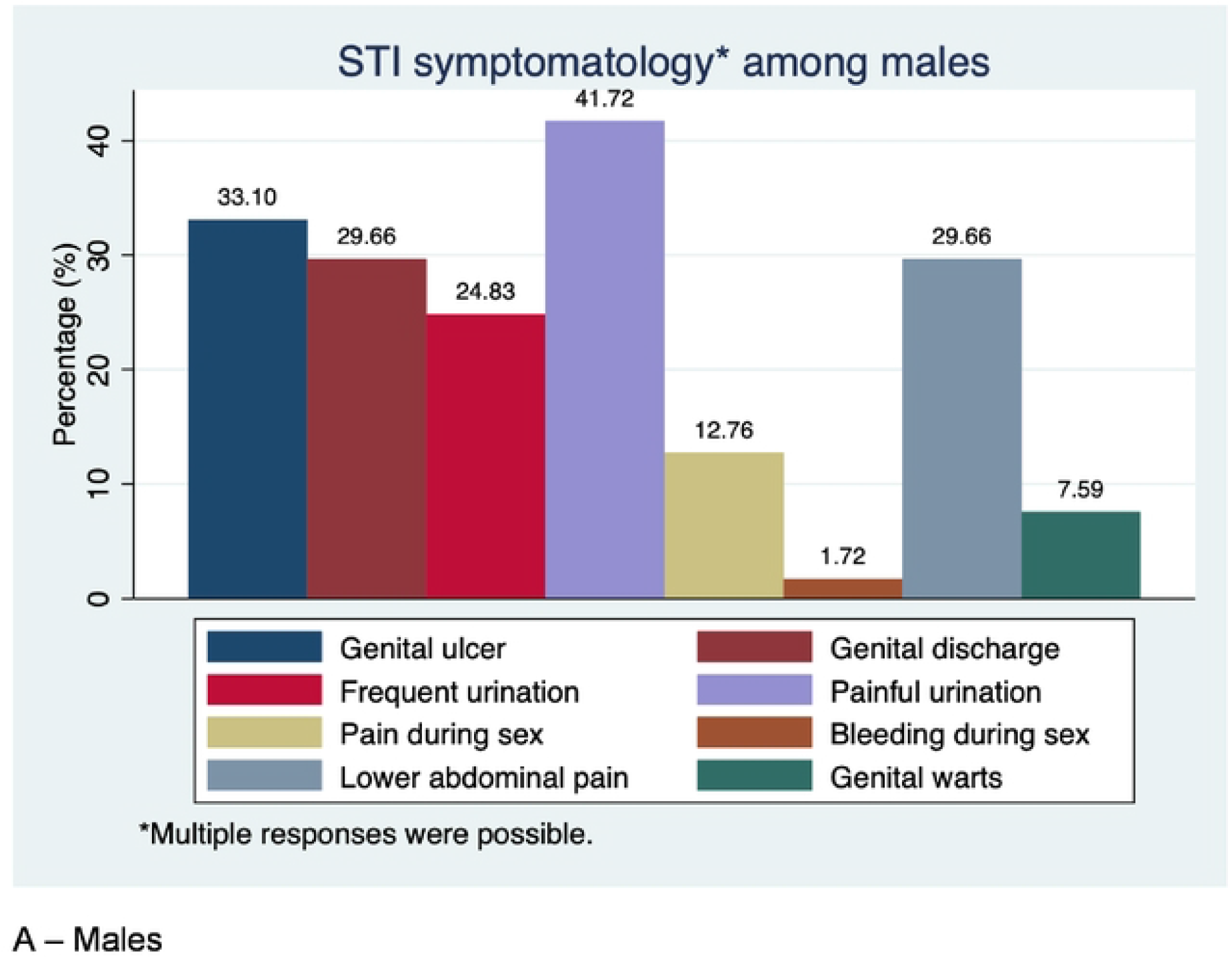
Symptomatology of STIPS participants who reported STI-related symptoms in past 6 months (N=962), by gender

### Treatment-seeking for STI symptoms

Overall, 545 participants (57%) reported seeking any treatment for their symptoms, while the remaining 43% reported seeking no treatment at all. The likelihood of seeking any treatment was similar between men and women (160/290 [55%] males; 385/671 [57%] females; PRR: 1.04, 95% CI: 0.92-1.18). The prevalence of seeking any treatment also did not differ by community type (PRR: 1.04, 95% CI: 0.93-1.16). No significant differences were observed when further stratifying by gender and community type.

Figure 2 shows where participants sought treatment for their symptoms. Nearly all participants (95%, 515/541) who sought treatment did so at only one location, while 5% (26/541) reported seeking treatment at two locations. Of those who sought treatment, 58% sought treatment at a private clinic, 28% at a government clinic, 9% at a pharmacy/drug store, 3% at a traditional healer, 2% at a market/shop, and 5% at some other location. Private clinics were the most common treatment location among both genders (71% among males; 53% among females). RHSP clinics comprised 7% of all private clinic visits. Comparing private and government clinics, women were less likely to seek treatment at private clinics (more likely to seek government clinics) than men (PRR: 0.76, 95% CI: 0.68-0.85). This trend was seen in both the fishing (PRR: 0.86, 95% CI: 0.76-0.96) and inland (PRR: 0.65, 95% CI: 0.51-0.83) communities. When comparing men across communities, men in fishing communities were significantly more likely to seek treatment at a private clinic than men in inland communities (PRR: 1.25, 95% CI: 1.04-1.51). Similarly, women in the fishing community were significantly more likely to seek treatment at private clinics than women in the inland community (PRR: 1.65, 95% CI: 1.36-2.01).

**Fig 2.**
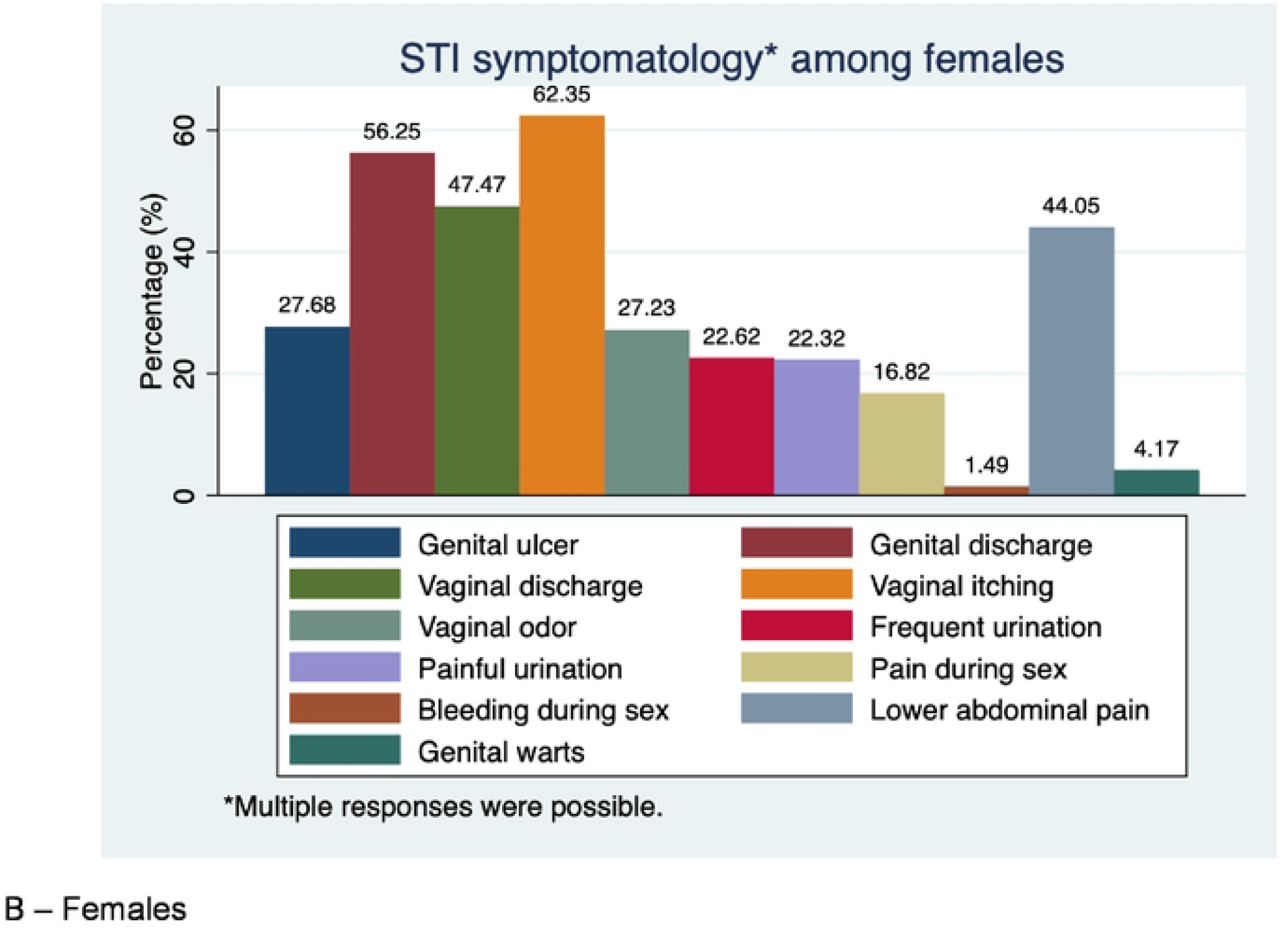
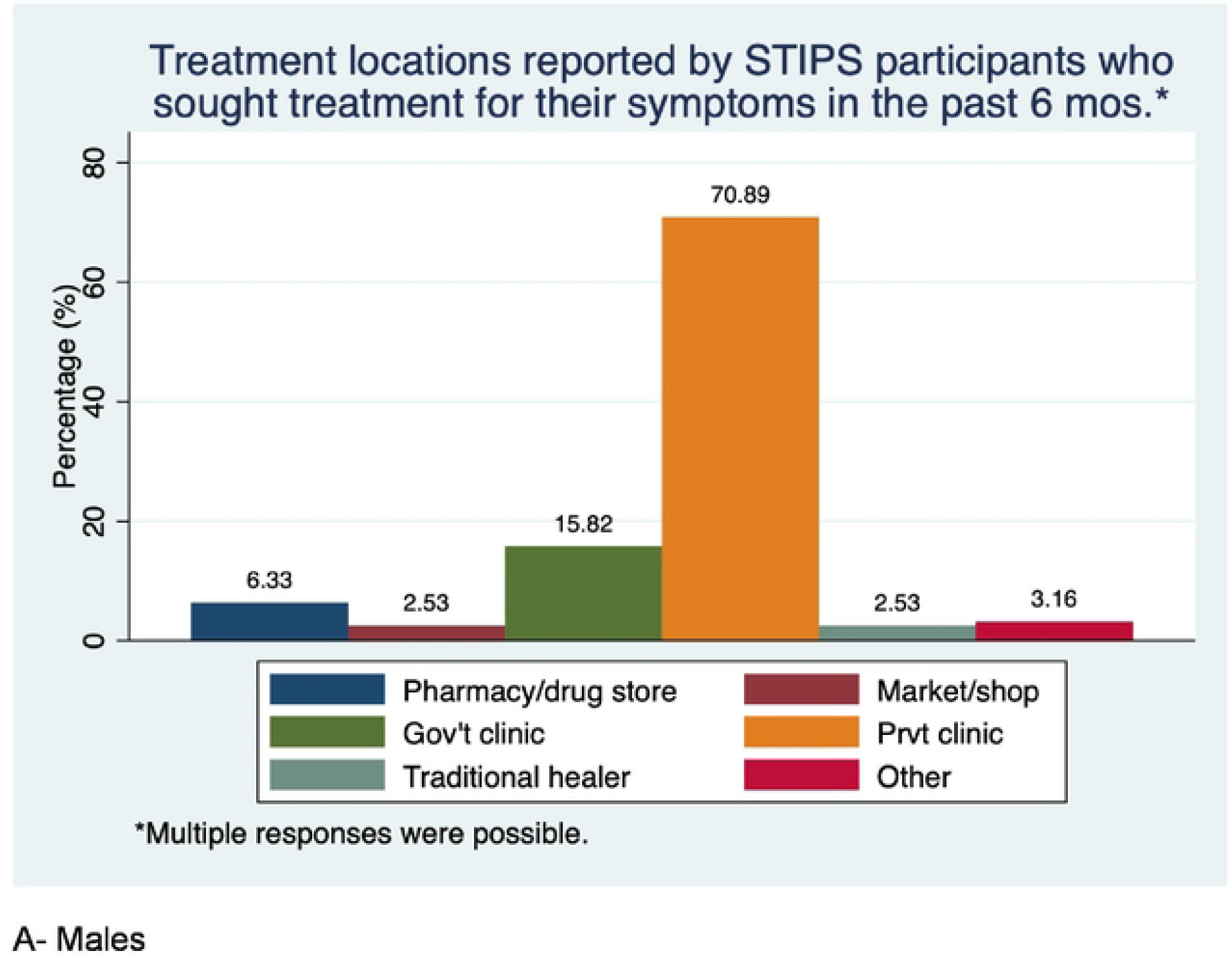
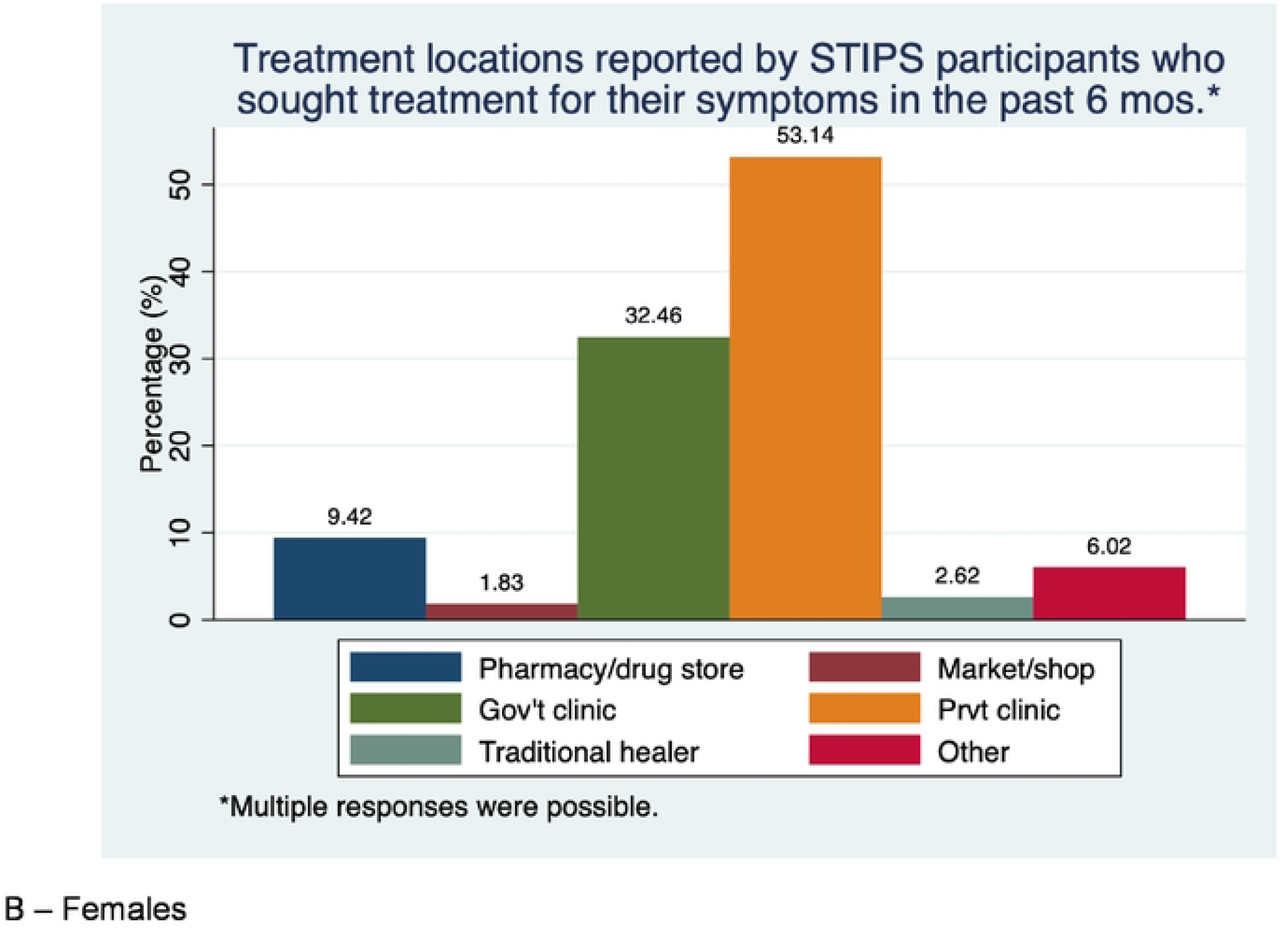
Treatment locations reported by STIPS participants who sought treatment for their symptoms in the past 6 months (N=545)

### Treatment-seeking at a clinic

A total of 457 participants (48%) reported seeking treatment at a clinic (government or private) for their symptoms. There was no difference in the prevalence of clinic treatment by gender (47% males; 48% females; PRR: 1.02, 95% CI: 0.88-1.18) or by community type (48% inland; 47% fishing; PRR: 0.98, 95% CI: 0.78-0.86) When stratifying each gender by community type, however, we found that men in the fishing community were significantly less likely to seek clinic treatment than men in the inland community (55% men in inland; 43% men in fishing; PRR: 0.78, 95% CI: 0.61-1.00).

Descriptive data of clinic treatment-seeking by gender is presented in Table 2; descriptive data of clinic treatment-seeking for the full sample is presented in Supplementary Table 1. Table. Overall, of men who sought clinic treatment, most were aged 20-39 years, married in a monogamous union, and working in the fishing industry. Of women who sought clinic treatment, most were aged 20-29 years, married in a monogamous union, and engaged in agriculture or housework. Thirty-three percent of men and 35% of women who sought treatment at a clinic were HIV-positive. With respect to STI symptomatology, painful urination (52%), genital discharge (47%) and genital ulcers (35%) were the most common symptoms reported among men who sought treatment at a clinic. Vaginal itching (69%), genital discharge (57%) and vaginal discharge (50%) were the most frequently reported symptoms among women who sought clinic treatment. About half of men who reported seeking treatment at a clinic reported 2-4 STI symptoms in the past six months (73/136 [54%]). This was similar for women (157/321 [49%]).

**Table 2.**
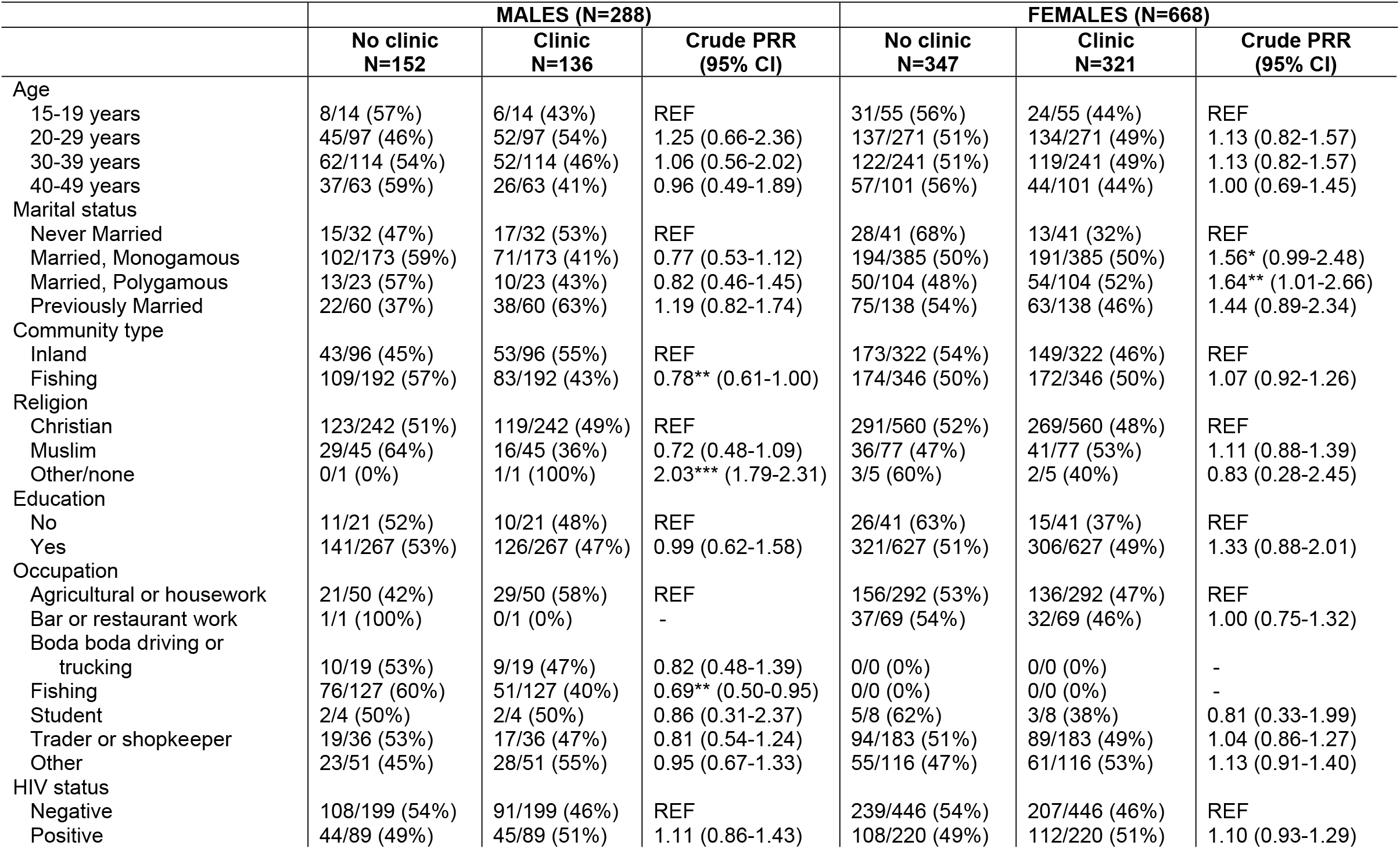
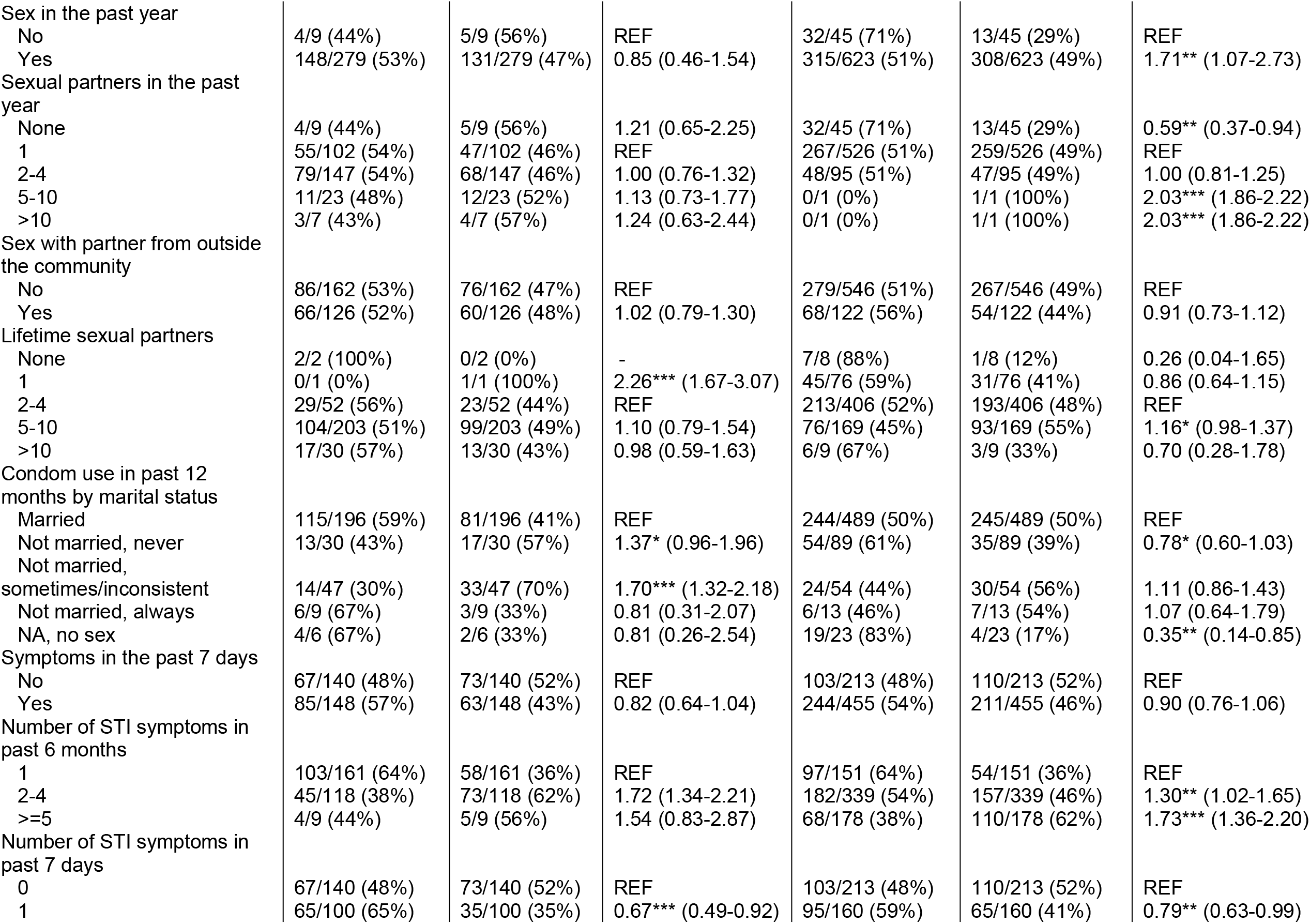
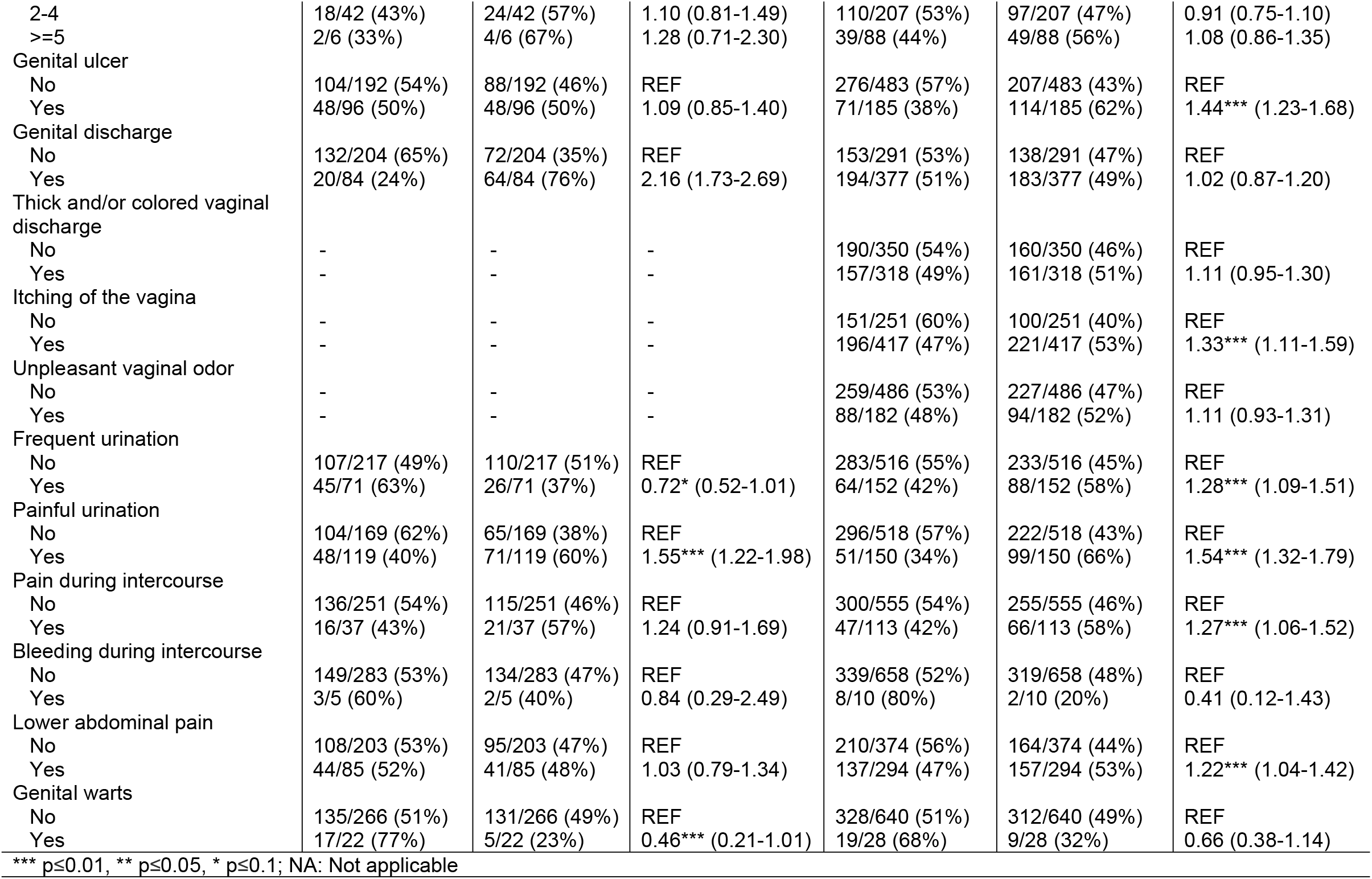
Unadjusted prevalence of clinic treatment-seeking among STIPS participants who reported STI symptoms in the past 6 months (N=956), by gender. Data are presented as n (%).

Just over half of men who reported seeking care at a clinic for their symptoms reported no symptoms in the past week (73/136 [54%]). In contrast, the majority of women who reported seeking care at a clinic for their symptoms reported having at least one STI symptom in the past week (66%), with 65/321 [20%] reporting one symptom 97/321, [30%] reporting 2-4 symptoms and 49/321 [15%] reporting five or more symptoms. When considering only those who reported at least one symptom in the past week, we saw that, for both men and women, just over half reported that they did not seek treatment at a clinic in the past six months for their symptoms (85/148 [57%] and 244/455 [54%], respectively). We found no significant differences in any of these associations in stratifications by both gender and community type.

Table 2 describes the univariable associations between sociodemographic characteristics, sexual behaviors, and STI symptomatology with clinic treatment for the full sample, as well as for men and women. Women were more likely to seek clinic treatment if they were in a polygamous marriage (PRR: 1.64, 95% CI: 1.01-2.66) and less likely to seek treatment if they had no sexual partners in the past year (PRR: 0.59, 95% CI: 0.37-0.94). Condom use, the number of STI symptoms reported in the past six months and the number of STI symptoms reported in the past week were also associated with clinic treatment-seeking for both men and women. Certain symptoms also showed univariable associations with clinic treatment-seeking for each gender. For men, the likelihood of seeking treatment in a clinic increased with reporting of painful urination in the past six months (PRR: 1.55, 95% CI: 1.22-1.98) and more than doubled with reporting of genital discharge (PRR: 2.16, 95% CI: 1.73-2.69). However, the likelihood for seeking clinic care decreased with reporting of genital warts (PRR: 0.46, 95% CI: 0.21-1.01). For women, the likelihood of seeking care at a clinic increased with reporting of genital ulcer (PRR: 1.44, 95% CI: 1.23-1.68), vaginal itching (PRR: 1.33, 95% CI: 1.11-1.59) frequent urination (PRR: 1.28, 95% CI: 1.09-1.51), painful urination (PRR: 1.54, 95% CI: 1.32-1.79), pain during intercourse (PRR: 1.27, 95% CI: 1.06-1.52) and lower abdominal pain (PRR: 1.22, 95% CI: 1.04-1.42) in the past six months. We found no significant differences in any of these associations in analyses stratified by community type.

In multivariable analyses, significant factors associated with seeking treatment at a clinic for men included being from the inland community and having multiple STI-related symptoms in the past six months. For women, the only significant factor associated with seeking STI treatment at a clinic was having multiple STI-related symptoms in the past six months (Table 3).

**Table 3.** Predictors of clinic treatment-seeking among STIPS participants who reported STI symptoms in the past 6 months (N=956), by gender

|  | MALES | FEMALES |
| --- | --- | --- |
|  | Adjusted PRR<br>(95% CI) | Adjusted PRR<br>(95% CI) |
| Age |  |  |
| 15-19 years | REF | REF |
| 20-29 years | 1.10 (0.61-2.00) | 0.97 (0.70-1.33) |
| 30-39 years | 1.01 (0.54-1.90) | 0.96 (0.69-1.33) |
| 40-49 years | 0.85 (0.44-1.64) | 0.89 (0.61-1.31) |
| Community type |  |  |
| Inland | REF | REF |
| Fishing | 0.70*** (0.55-0.89) | 0.97 (0.82-1.15) |
| Marital status |  |  |
| Never Married | REF | REF |
| Married, Monogamous | 0.92 (0.61-1.37) | 1.57* (0.96-2.58) |
| Married, Polygamous | 0.91 (0.52-1.59) | 1.67* (0.99-2.81) |
| Previously Married | 1.44* (0.97-2.14) | 1.46 (0.86-2.47) |
| HIV status |  |  |
| Negative | REF | REF |
| Positive | 1.27* (0.98-1.67) | 1.08 (0.90-1.30) |
| Number of STI symptoms in past 6 months |  |  |
| 1 | REF | REF |
| >1 | 1.73*** (1.36-2.21) | 1.41*** (1.12-1.78) |

\*\*\* p≤0.01, \*\* p≤0.05, \* p≤0.1

### Previous treatment-seeking and current STI prevalence

Among those who reported STI-related symptoms, CT prevalence was 11%, NG was 10%, TV was 13% and active syphilis was 7% When we restricted our analysis to only those individuals who reported previously seeking clinic treatment for their symptoms, we found that approximately one-third tested positive for any curable STI (CT, NG, TV or active syphilis) at the time of the survey (45/136 [33%] males; 98/321 [31%] females) (Table 4). We found no significant difference in the current prevalence of curable STIs between those who did and did not previously seek clinic treatment, for either gender (for men: 33% with curable STI who previously sought treatment versus 27% with curable STI who previously did not seek treatment; PRR: 1.23, 95% CI: 0.86-1.75); for women, 31% with curable STI who previously sought treatment versus 33% with curable STI who previously did not seek treatment; PRR: 0.92, 95% CI: 0.74-1.15).

**Table 4.**
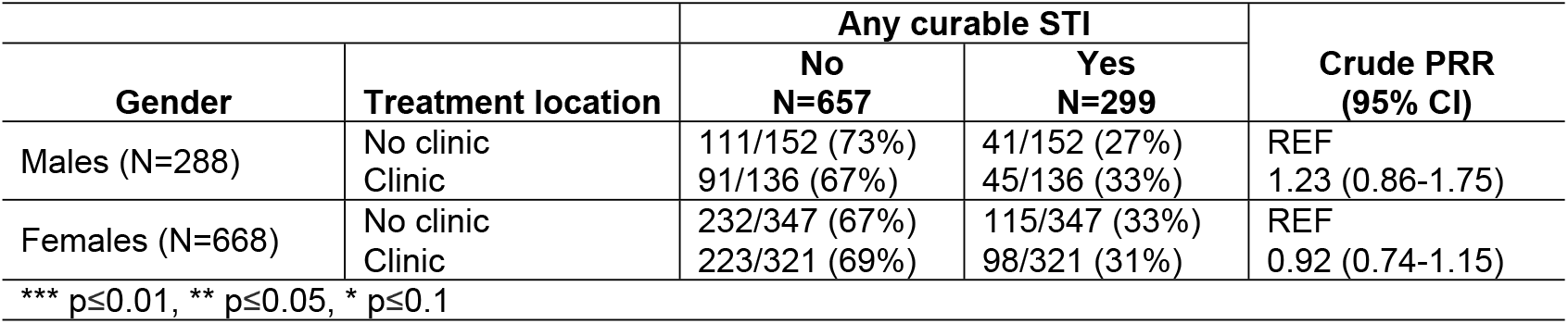
Prevalence of any curable STI (CT, NG, TV or active syphilis) at the time of the survey among STIPS participants who reported STI symptoms in the past 6 months (N=956), by gender

## Discussion

This study provides a population-based assessment of STI-related symptoms and treatment-seeking in rural Uganda and assesses factors associated with seeking treatment at a government or private clinic. Just over half (57%) of adults who reported STI-related symptoms in the past six months reported seeking any treatment for their symptoms, with similar rates in men and women. Seeking treatment at a clinic was 48% overall (47% among men; 48% among women). While our estimates were based on a population-based sampling approach of all eligible adults in our study communities, our estimations of clinic treatment-seeking are lower than those found in previous studies, including the 2016 Ugandan DHS (70% for the country) and the first round of the RCCS in the 1990s (67% for the region) [5, 20].

Our data showed no difference in the prevalence of clinic treatment-seeking by gender. This was unexpected, as research in Uganda and other resource-limited settings suggest gendered patterns of treatment-seeking [11, 21–27], with women more likely to delay, and less likely to seek, STI treatment than men [28–31]. While the 2016 Uganda DHS estimates show a higher prevalence of clinic treatment-seeking among women than men [5], as aforementioned, we are hesitant to compare our findings to DHS estimates because of urban/rural differences. We recommend that future researchers continue to assess gender-specific treatment-seeking behavior across a range of settings.

We also found that men in the fishing community were significantly less likely to seek clinic treatment, as compared to men in the inland community. The epidemiologic differences between fishing and inland communities in Rakai are well-established: fishing communities show a disproportionate burden of HIV, high prevalence of sexual risk behaviors and historically showed a lower use of combination HIV prevention services (though this has been increasing in recent years with significant new service provision) [32–35]. Furthermore, data show that overall, men in Rakai are less likely to be enrolled in HIV care [36, 37], as are in-migrants [36]. Rakai fishing communities, or ‘landing sites’, generally have a high proportion of men, the majority of whom migrate from other communities in order to work as fishermen on Lake Victoria. Assuming that the barriers to HIV care for Rakai residents – including stigma, demanding work schedules, transport costs, belief in spiritual healing, long wait times and inadequate staff respect for patients [38] – also apply to STI treatment-seeking, our observations indicating a lower prevalence of clinic treatment-seeking among men in the fishing community is not unexpected. What is surprising, however, is the fact that this difference was not observed among women. We recommend future researchers explore the intersection of gender and community type, as well as try to better understand the barriers to treatment-seeking in fishing communities among men.

In addition, we found the presence of some, but not all, symptoms to be associated with clinic treatment-seeking for each gender. For instance, lower abdominal pain, pain or bleeding during intercourse, and genital ulcers showed no association with treatment-seeking among men in our study. The same goes for genital warts and thick/colored discharge among women. Not recognizing STI-related symptoms, not perceiving them as severe, or not attributing them to STI-related causes can prevent treatment-seeking [1, 14, 39, 40]. A similar phenomenon may have occurred in our sample: participants may not have attributed lower abdominal pain, warts or discharge to an STI, thereby explaining why the presence of some symptoms were associated with clinic treatment over others.

The frequency of private clinic use in our sample was notable. The Ugandan healthcare system suffered losses during the decades of civil unrest in the 1980s [41]. Consequentially, many Ugandans have come to perceive health centers as expensive and lacking medication, and often turn to self-medication first and use health centers as a last resort [41–45]. Also as a result of the political turmoil, the number of public health services in the Uganda decreased and the number of private clinics increased [45, 46]. While we did not include a treatment location mapping exercise in our study, we did find a 2010 study that mapped the availability of private and public facilities in rural areas of Uganda [47]. Based on their work, the authors reported that public facilities made up 4.3% of all the health care units that were mapped as compared to private facilities which made up 95.7%. Private-for-profit clinics and drug shops made up 17.1% of all mapped facilities and private-not-for-profit facilities made up 1.6% of all mapped facilities [47]. While still considered rural, the districts included in their study are more central and developed than Rakai district. Nevertheless, we expect that their finding of more private than public clinics may still apply to Rakai. As such, the high prevalence of private clinic treatment that we observed may be partially explained by the high availability of private clinics in the area.

We also found a difference in private versus government clinic treatment-seeking by gender, with women more likely to attend government clinics than men as seen in other settings [48–50]. A lack of finances, unfriendly reception and long wait times have been identified as reasons for why women do not seek care at formal sector clinics [50].

The lack of association between clinic treatment-seeking and HIV status surprised us; we expected that individuals who were HIV-seropositive to be more likely to seek clinic treatment for their symptoms than those who were HIV-negative. This was observed among Rwandan women [51]. Eighty-six percent of PLHIV in our study reported currently taking antiretroviral treatment (ART); we would have expected that their routine interaction with the health system due to ART would make them more likely to attend a clinic for treatment than those without HIV. PLHIV may also be more conscious of their sexual risk behavior and sexual health than those who are negative [52–54], furthering their likelihood of seeking clinic treatment. We speculate that the availability and affordability of services or stigma may have affected treatment-seeking behavior among PLHIV and recommend further research in this area.

We also found that approximately one-third of men and women who previously sought clinic treatment for their symptoms were diagnosed with at least one curable STI (CT, NG, TV or active syphilis) at the time of the survey. Furthermore, our analysis showed no difference in the current prevalence of curable STIs comparing those who previously sought clinic treatment versus those who did not, for either gender. Assuming that seeking clinic treatment meant receiving treatment, these data could indicate that reinfection rates were high, treatment was inadequate, or both. Further studies exploring the temporal association between past treatment-seeking, including receiving and adhering to treatment, and current STI prevalence are recommended to assess treatment effectiveness.

A strength of our study lies in its population-based sample, which is rare in other studies focused on treatment-seeking behavior. Calls have been made for a broader research perspective in order to understand sexual healthcare seeking behavior [55]. This perspective includes a focus on non-attendance at healthcare services as well as research that uses non-patient samples recruited from non-medical settings in order to accurately capture the range of behaviors, perspectives, and health issues occurring within the population and ensure appropriate and effective service provision [34]. We addressed these items in our study by interviewing all eligible individuals in our study communities and including both persons who did and did not seek treatment. Taken together, this information can provide program managers and decisionmakers with a better understanding of treatment-seeking patterns within the community.

However, our research is not without limitations. First, our estimations of clinic treatment-seeking may be lower than national estimates because of the communities included in our sample. The communities that we included are considered rural and treatment-seeking tends to be lower in rural settings [1, 28, 56]. Second, we did not further define treatment settings or ask participants to name specific treatment locations. As a result, some treatment locations may have been misclassified (e.g. drug shops being reported as private clinics, etc.). Third, inclusion of study participants was based on self-reported symptom history in the past six months. It is possible that eligible participants may not have been included because they were too shy, or embarrassed, to share their symptom history (social desirability bias), did not remember their symptoms (recall bias) or did not understand the terms we used for the symptoms. Fourth, given that the survey was administered by RHSP staff, it is possible that participants over-reported seeking clinical care or under-reported seeking care in informal sectors (or not seeking any care at all) in order to please the interviewer. Given the lower than expected rates of treatment-seeking, however, we doubt this was a major concern. Fifth, our study did not assess environmental, social, psychosocial, economic, geographic or service-related factors, as well as symptom severity, which have been shown to be associated with treatment-seeking behavior in low-resource settings [1]. We also did not measure the availability of each type of treatment location in our study communities – this information would be useful to contextualize our results. Sixth, we note that, while our study was sufficiently powered to assess differences between men and women, we may have been underpowered to detect differences by both gender and community type. This sub-analysis may be of programmatic interest; we recommend that researchers consider this when designing future studies.

Finally, our analysis grouped together private and government clinics, and compared them to other treatment locations/no treatment. In doing so, we assumed that these clinics provided appropriate and effective care, of equal/sufficient quality, and that anything but clinical care was ineffective. This may be an unfair assumption: in low-resource settings, formal sector facilities often show poor quality of care in general [57, 58]. A study specifically on the quality of STI case management in Ugandan private clinics and drug shops concluded that the quality of management was poor [41]. We recommend studying quality of care and barriers to providing quality care in local facilities, as well as urge leadership to strengthen care and enforce quality standards across health service sectors and facilities.

## Conclusion

Timely and appropriate diagnosis is critical to STI treatment and control. We found that half of adults with STI symptoms in two rural Ugandan communities are not seeking appropriate clinical care under the syndromic management strategy. These individuals are a priority for public health intervention. We recommend researchers continue to focus on treatment-seeking behavior in low-resource settings and explore barriers to seeking care, including health system barriers such as low quality of care; we urge decision-makers to increase support for STI services in this and similar contexts.

## Data Availability

A deidentified version of the Rakai Community Cohort Study data can be provided to interested parties subject to completion of the Rakai Health Sciences Program data request form and signing of a Data Transfer Agreement. Inquiries should be directed to.

## Acknowledgements

We are grateful to the community members and leaders of Rakai who participated in this research. We also give special appreciation to all the RHSP staff members and in particular the STIPS team for supporting this research. We would also like to acknowledge the support of Drs. Ronald Gray and Maria Wawer for their support for this project undertaken at Rakai, and Joyce Yehjin Jang and Ping Teresa Yeh for their help preparing this manuscript for submission.

